# Effects of cash transfer and incentive programs on service utilization and treatment outcomes related to neglected tropical diseases and their impact on health and nutrition in low and middle-income countries: a protocol for a systematic review

**DOI:** 10.1101/2025.03.18.25324206

**Authors:** S. M. Tafsir Hasan, Radhika Dayal, Amena Al Nishan, Sunetra Ghatak, Puja Chakraborty, Sudeshna Maitra, Sumaiya Tasneem Raisa, Nizamuddin Khan, Dinesh Mondal, Tahmeed Ahmed, Avishek Hazra

## Abstract

**Background:** Although there has been a general decrease in the global burden on Neglected tropical diseases (NTDs), progress is not on track to meet the Sustainable Development Goals 2030. Evidence suggests the positive effect of conditional cash transfer programs on controlling NTDs. However, it is to be evaluated whether other financial incentive programs exert similar effects on NTD-related service utilization and treatment outcomes and, in turn, on health and nutrition in low and middle-income countries (LMICs). This proposed systematic review aims to evaluate the effects of cash transfer and incentive programs on NTD-related service utilization and treatment outcomes in LMICs, to examine how covariates, program design, implementation plan, and incentive program design influence these outcomes, and to examine how NTD-related these outcomes influence nutritional status and health in LMICs.

**Methods:** This systematic review will follow the PICOS (Population, Interventions, Comparators, Outcomes, Studies) framework. The population of this review will be restricted to the adult population and children of all ages of LMICs. For intervention selection, we will include any program or policies addressing socioeconomic disadvantage through the provision of cash transfers or incentive programs, including but not limited to in-kind transfers, food vouchers, free medicine vouchers, discount coupons, and micro-credit to households or individuals. Eligible comparators will be the population who did not receive any intervention, recipients of interventions from before the intervention, and populations or areas exposed to different levels of intervention coverage. The primary outcomes of interest will be service utilization and treatment outcomes related to any of the 21 NTDs and disease groups prioritized by WHO. Secondary outcomes include variables related to general health and nutrition outcomes.

**Discussion:** To provide a comprehensive and unbiased assessment of the current state of knowledge on the effects of cash transfer and other financial incentive programs on NTD-related service utilization and treatment outcomes in LMICs, conducting a systematic review is crucial. This systematic review will provide comprehensive, and unbiased evidence considering the strengths and limitations of existing knowledge on NTDs. Additionally, we will also explore how incentive type and program design influence NTD-related service utilization and treatment outcomes in LMICs.

**Ethics and dissemination:** The systematic review protocol has been reviewed and approved by two independent institutional review boards (IRBs): icddr,b (PR-24112) in Bangladesh and Population Council Institute (IORG0011824) in India. The results will be submitted to a peer-reviewed journal and presented at national and international conferences to reach the scientific community.

**Systematic review registration:** The protocol has been registered in PROSPERO (CRD42024627804).

## INTRODUCTION

Neglected Tropical Diseases (NTDs) remain a major global public health concern, affecting an estimated 1.62 billion people in tropical and subtropical climatic zones (1, 2) and accounting for approximately 200,000 deaths and 14.5 million disability-adjusted life years (DALYs) annually (2, 3). Although there has been a general decline in the global burden of NTDs (2), the progress is not yet on track to meet the ambitious Sustainable Development Goals (SDGs) 2030 target of a 90 percent reduction in the number of people requiring interventions for NTDs (3). NTDs continue to pose a big challenge for low- and middle-income countries (LMICs) due to their intricate links with poverty, inadequate hygiene, poor sanitation, and limited healthcare access (4, 5).

Poverty is a key factor exacerbating the burden of NTDs in LMICs. Financial deprivation and disease are closely intertwined, which influence the social and environmental determinants of NTDs, including water, sanitation, and hygiene (WASH), education, housing, and healthcare accessibility (5–7). Poverty highly modulates the accessibility and utilization of the services related to the five important NTD control strategies recommended by the World Health Organization (WHO): innovative and intensified disease management, preventive chemotherapy, vector control, veterinary public health measures, and provision of safe WASH practices (8, 9). NTDs disproportionately affect impoverished and marginalized communities, resulting in increased catastrophic health spending (CHS) and out-of-pocket (OOP) expenses (10). Already living in poverty before the onset of these diseases, affected households often face deeper financial hardship as rising OOP costs push them further below the poverty line (11–13). To manage these expenses, they often resort to coping strategies such as borrowing money or selling assets, which further exacerbates their poverty. This cycle undermines access to healthcare services and negatively affects treatment outcomes (13). Physical disabilities due to NTDs, such as blindness, lesions, and swelling, may also contribute to decreased income ability (14). This financial burden perpetuates a cycle of poverty and NTD burden, hindering service uptakes and treatments (15). Consequently, the detrimental effects of NTDs are amplified, including poor NTD-related service utilization and adherence, treatment failure, disability, and deterioration of general health and well-being (15).

Moreover, infection with NTDs increases the nutritional burden of the hosts and impedes overall health. NTDs have been reported to be associated with stunting, anemia, iron deficiency, micronutrient deficiencies, and malnutrition (16–19). Simultaneously, nutrient deficiency may also increase the susceptibility and vulnerability of NTD-affected individuals and lead to NTD treatment failure (18, 20, 21). Actions taken to control NTDs may eventually improve nutritional outcomes which, in turn, will also influence overall health and wellbeing, particularly in resource-limited settings.

Despite various programs and policies implementing innovative strategies to improve NTD campaign coverage, multiple layers of poverty-associated and financial obstacles continue to hinder appropriate health-seeking behaviors in affected populations. These barriers include attending screening clinics, receiving adequate care, and treatment adherence (22–25). Cash transfer programs have demonstrated potential for improving NTD outcomes (26–29). These programs aim to provide small-scale monetary assistance to individuals or households to break the intergenerational cycle of poverty. Conditional cash transfers (CCT) impart benefits through a two-way strategy: direct cash assistance and enforced conditionalities (30). It is also possible that conditionalities related to the control of NTDs may also impart positive effects on nutrition and overall health outcomes in LMICs. Large-scale, national-level cash transfer programs in Latin America have not only proven effective in improving social protection, economic stability, and sustained developmental outcomes (31) but have also generated interest in using cash transfers to target context-specific health outcomes, including NTD treatment outcomes, maternal health, child health, cognitive development, nutrition, and healthcare utilization (26, 32–34).

A recent systematic review highlighted the positive impact of conditional cash transfer programs in controlling NTDs. It indicated that the targeted benefits of these programs for vulnerable populations can help reduce health inequalities associated with NTDs. For example, cash transfers have been shown to reduce leprosy incidence and increase the uptake of deworming treatments in certain contexts (35). However, the review had a major limitation in that it included and synthesized evidence for only 3 of the 21 neglected tropical diseases and disease groups prioritized by WHO (2, 3, 35). While other incentive programs, such as, in-kind transfers, micro-credits, vouchers for transport, medicines, or food, have shown the potential to promote health knowledge, reduce health inequalities, and improve health-seeking behaviors, their direct or indirect effects on NTD-related indicators have not been comprehensively studied in LMICs (36, 37).

Given this context, there is a need for an evidence synthesis on the effects of cash transfers and other incentive programs on NTD-related service utilization and treatment outcomes in LMICs. This proposed systematic review and meta-analysis (depending on information availability), will conduct a comprehensive synthesis and unbiased assessment of the existing studies in this area, critically assessing the strengths and limitations of existing evidence.

### Objectives

The systematic review aims to answer the following three questions: 1) What are the effects of cash transfer and incentive programs on NTD-related service utilization and treatment outcomes in LMICs? 2) How do program design, implementation plan, and other covariates such as incentive program design (type, amount, and frequency) influence NTD-related service utilization and treatment outcomes in LMICs? 3) How do NTD-related service utilization and treatment outcomes influence nutritional status, overall health, and well-being in LMICs?

## METHODS

This protocol was developed in accordance with the guidelines of the Preferred Reporting Items for Systematic Reviews and Meta-Analyses for Protocols (PRISMA-P) (38). A PRISMA-P checklist for this protocol is provided in Supplemental File 1. The protocol has been registered in PROSPERO (CRD42024627804).

### Eligibility criteria

The eligibility criteria for studies to be included in this review are adapted based on the PICOS framework (Population, Interventions, Comparators, Outcomes, Study Designs) (39), as described below.

#### Population

The population of interest in this review will be adults and children of all ages residing in LMICs.

#### Interventions

For intervention, this review will include programs or policies addressing socioeconomic disadvantages through the provision of cash transfers or financial incentive programs, including but not limited to in-kind transfers, food vouchers, free medicine vouchers, discount coupons, and micro-credit to households or individuals.

CCT programs require beneficiaries to comply with certain conditionalities (e.g., regular health check-ups) (40), while unconditional cash transfer programs do not set such requirements (41). Microcredit is the extension of very small loans (microloans) to impoverished borrowers who typically lack collateral, steady employment, and a verifiable credit history (42). It is designed to support entrepreneurship and alleviate poverty (42). An in-kind transfer is a transfer of goods or services rather than cash (43). Other incentive programs may also include giving free medicine vouchers, food vouchers, discount coupons, social protection, social programs, scholarship programs, family allowances, and micro-finance.

#### Comparators

This review will include studies with suitable comparators; studies without comparators will be excluded. Comparators will include individuals or households who did not receive any cash transfers or incentive programs, recipients of cash transfers or incentive programs from pre-intervention period, and populations or areas with varying levels of intervention coverage.

#### Outcomes

The primary outcomes of interest will be service utilization and treatment outcomes related to any of the 21 NTDs and disease groups prioritized by WHO (2). These include Buruli ulcer; Chagas disease; dengue and chikungunya; dracunculiasis; echinococcosis; foodborne trematodiases; human African trypanosomiasis; leishmaniasis; leprosy; lymphatic filariasis; mycetoma, chromoblastomycosis and other deep mycoses; noma; onchocerciasis; rabies; scabies and other ectoparasitoses; schistosomiasis; soil-transmitted helminthiases; snakebite envenoming; taeniasis/cysticercosis; trachoma; and yaws.

Service utilization related to WHO-recommended core strategic interventions for each of the 21 NTDs and disease groups will be evaluated. The aspects of service utilization evaluated will include, but not be limited to, health-seeking behavior, attendance at screening clinics, treatment adherence, and NTD campaign coverage. A comprehensive list of core strategic interventions and treatment outcomes for each of the 21 NTDs and disease groups is provided in Table 1 (1, 3, 44–49).

**Table 1.** List of core strategic interventions and treatment outcomes for each of the 21 NTDs and disease groups.

| Serial | NTD name | Core strategic interventions |  |  |  |  | Treatment outcomes |
| --- | --- | --- | --- | --- | --- | --- | --- |
|  |  | Innovative and intensified disease management | Preventive chemotherapy | Vector control | Veterinary public health measures | Provision of safe WASH practices |  |
| 1 | Buruli ulcer | Early detection, case management (provision of antibiotics: rifampicin, clarithromycin, moxifloxacin, streptomycin, and hygienic wound care) | Bacillus Calmette–Guérin (BCG) vaccination | Aquatic insects, adult mosquitoes, possums, and biting arthropods | Not applicable | Safe water storage practices | Disability, morbidity management including surgery, wound, and lymphoedema management, physiotherapy and long-term rehabilitation, prevalence, incidence, detection rate, cure rate, disability rate, death rate |
|  |  | Innovative and intensified disease management | Preventive chemotherapy | Vector control | Veterinary public health measures | Provision of safe WASH practices |  |
| 2 | Chagas disease | Case management (benznidazole and nifurtimox), mass drug administration, surgical treatment | Blood screening before blood transfusion and organ transplantation | Spraying with residual insecticides to remove triatomine bugs, home cleanliness, and housing improvements (e.g. crack-free walls, bed nets), wear protective clothing | Not applicable | Good hygiene practices | Prevalence, incidence, detection rate, cure rate, surgery rate, disability rate, death rate |
| 3 | Dengue and chikungunya | Case management (fluid management, proactive treatment of hemorrhage, such as platelet transfusion or whole blood transfusion, paracetamol) | Vaccination (dengue) | Reducing mosquito habitats (e.g. environmental modification, use of lids), wire mesh, mosquito | Not applicable | Safe water storage practices, safe water disposal, waste management | Prevalence, incidence, detection rate, cure rate, disability rate, death rate |
|  |  | Innovative and intensified disease management | Preventive chemotherapy | Vector control | Veterinary public health measures | Provision of safe WASH practices |  |
|  |  |  |  | net, mosquito trap, mosquito insecticide, mosquito repellents |  |  |  |
| 4 | Dracunculiasis | Surveillance, early detection of cases, case containment, cash reward scheme for voluntary reporting, metronidazole or thiabendazole, health education | Not applicable | Temephos to control water fleas/cyclops, | Proactive tethering of dogs | Safe drinking water supply and sanitation, water filtration, preventing water contamination | Prevalence, incidence, detection rate, cure rate, disability rate, death rate |
| 5 | Echinococcosis | Albendazole prophylaxis, surgery, early detection by ultrasound | Not applicable | Not applicable | Deworming of dogs, foxes, vaccination | Handwash, avoiding contaminated | Prevalence, incidence, detection rate, cure rate, disability rate, |
|  |  | Innovative and intensified disease management | Preventive chemotherapy | Vector control | Veterinary public health measures | Provision of safe WASH practices |  |
|  |  |  |  |  | of sheeps, safe slaughtering and disposal | food and water | surgery rate, total or partial cystopericytectomy, liver damage, death rate |
| 6 | Foodborne trematodiasis | Anthelmintic medicines (praziquantel, triclabendazole), PAIR (puncture, aspiration, instillation, and respiration) methods, surgery (partial hepatectomy), Provision of care and rehabilitative services | Mass drug administration with triclabendazole (*Fasciola* spp.) and praziquantel (small liver flukes and *Paragonimus* spp.) | Not applicable | Livestock and domestic animal treatment, safe fish farming, one health intervention | Safe disposal of faecal waste, safe water, safe food practices, safe WASH practices | Prevalence, incidence, detection rate, cure rate, disability rate, surgery rate, liver damage, death rate |
| 7 | Human African trypanosomiasis | Screening clinic attendance, active case detection, treatment of gHAT: pentamidine, | Not applicable | Baits and traps, impregnated screens, insecticide | Treatment of animals (cattle, pigs) | Safe water, safe WASH practices | Prevalence, incidence, detection rate, cure rate, |
|  |  | Innovative and intensified disease management | Preventive chemotherapy | Vector control | Veterinary public health measures | Provision of safe WASH practices |  |
|  |  | eflornithine and nifurtimox, fexinidazole, melarsoprol, suramin |  | spraying, reducing the disease reservoir, controlling the tsetse fly vector |  |  | disability rate, death rate |
| 8 | Leishmaniasis | Early diagnosis, cryotherapy, thermotherapy, medication (liposomal amphotericin B, pentavalent antimonials, miltefosine), rapid diagnostic test rk-39 | Not applicable | Insecticide spraying, insecticide-treated nets, environmental management, protection from sandfly bites | Rodent control, vaccination for dogs | Safe WASH practices | Prevalence, incidence, detection rate, cure rate, disability rate, death rate |
| 9 | Leprosy | Early detection, mass drug administration, physical therapy, multidrug therapy (rifampicin, clofazimine, | Post exposure prophylaxis of rifampicin, vaccination with BCG, | Not applicable | Not applicable | WASH in health facility | Prevalence, incidence, detection rate, cure rate, disability rate, death rate |
|  |  | Innovative and intensified disease management | Preventive chemotherapy | Vector control | Veterinary public health measures | Provision of safe WASH practices |  |
|  |  | dapsone, ofloxacin, and minocycline) | Mycobacterium w (Mw) |  |  |  |  |
| 10 | Lymphatic filariasis | Essential package of care (skincare and hygiene, exercises for lymphoedema, treatment of adenolymphangitis), surgery to cure hydrocele, antifilarial medicines (albendazole, diethylcarbamazine citrate), ivermectin for lymphatic filariasis co-endemic countries | Mass drug administration (MDA), annual MDA with diethylcarbamazine citrate and albendazole, home-based disability alleviation and prevention | Vector control to reduce transmission, avoid mosquito bites | Not applicable | Hygiene, sanitation improvement, personal hygiene and self-care of the affected limbs to avoid secondary infections, agriculture, livestock, wildlife, environment (One Health), | Prevalence, incidence, detection rate, cure rate, disability rate, hydrocele management, surgery rate, mental health, death rate |
|  |  | Innovative and intensified disease management | Preventive chemotherapy | Vector control | Veterinary public health measures | Provision of safe WASH practices |  |
|  |  |  |  |  |  | avoid mosquito bites |  |
| 11 | Mycetoma, chromoblastomycosis and other deep mycoses (paracoccidioidomycosis/PCM, caused by *Paracoccidioides* spp. and sporotrichosis/ST, caused by *Sporothrix* spp.) | Long term itraconazole + antibiotic administration, physical therapy, wound care, surgery (excision, debridement, amputation) | Self-protection by wearing long clothes, closed-toed shoes | Not applicable | Pet control for sporotrichosis, feral cat management | Personal hygiene and self-care of the affected limbs to avoid secondary infections | Prevalence, incidence, detection rate, cure rate, disability rate, local excision, debridement, amputation, surgery rate, death rate |
| 12 | Noma | Case management with antibiotics, mouthwash, health education | Nutrition supplements | Not applicable | Not applicable | Personal hygiene | Prevalence, incidence, detection rate, cure rate, disability rate |
|  |  | Innovative and intensified disease management | Preventive chemotherapy | Vector control | Veterinary public health measures | Provision of safe WASH practices |  |
|  |  |  |  |  |  |  | (disfiguration), mental health, death rate |
| 13 | Onchocerciasis | Ivermectin, doxycycline, eye care, mass drug administration | Mass drug administration of ivermectin for community-based prevention | Insecticides at black fly habitats and larval breeding sites, avoid blackfly bites | Not applicable | Not applicable | Prevalence, incidence, detection rate, cure rate, disability rate (ocular morbidity rate), death rate |
|  |  | Innovative and intensified disease management | Preventive chemotherapy | Vector control | Veterinary public health measures | Provision of safe WASH practices |  |
| 14 | Rabies | Post-exposure prophylaxis (PEP) with rabies vaccine and rabies immunoglobulin | Pre-exposure vaccination for people at high risk (e.g. laboratory staff working with rabies virus, veterinarians and animal handlers) | Wild mammals (dogs, bats) control | Mass vaccination of dogs, mammal pet vaccination, agriculture, livestock, wildlife, environment (One Health) | Not applicable | Prevalence, incidence, detection rate, death rate |
| 15 | Scabies and other ectoparasitoses | Topical scabicides, permethrin, benzyl benzoate, malathion, and sulfur ointment, oral ivermectin | Mass drug administration (MDA) using oral ivermectin | Not applicable | Not applicable | WASH practices, hygiene practices | Prevalence, incidence, detection rate, cure rate, co-morbidities (kidney disease, rheumatic heart disease, systemic infection) |
|  |  | Innovative and intensified disease management | Preventive chemotherapy | Vector control | Veterinary public health measures | Provision of safe WASH practices |  |
| 16 | Schistosomiasis | Case management and surgery, education | Mass drug administration (MDA) with praziquantel | Snail control | Keeping animals away from transmission sites | Safe water, improved sanitation, excreta management, hygiene | Prevalence, incidence, detection rate, cure rate, disability rate, surgery rate, death rate |
| 17 | Soil-transmitted helminthiasis | Case management (with albendazole/ mebendazole/ ivermectin), education of hygiene | Mass drug administration (MDA) with albendazole/ mebendazole/ ivermectin, antenatal care preventive treatment | Not applicable | Not applicable | WASH practices, safe water, improved sanitation, excreta management, hygiene | Prevalence, incidence, detection rate, cure rate, disability rate, death rate |
|  |  | Innovative and intensified disease management | Preventive chemotherapy | Vector control | Veterinary public health measures | Provision of safe WASH practices |  |
| 18 | Snakebite envenoming | High-quality, safe, and effective snake antivenoms; wound care; ventilation; surgery; WHO Regional Action Plan for prevention and control of snakebite envenoming 2022–2030 | Protective footwear, sealing house wall, roof and door gaps, bed nets, household rodent controls to keep away the snakes | Not applicable | Not applicable | Not applicable | Prevalence, incidence, detection rate, cure rate, disability rate, surgery rate, Intensive Care Unit (ICU) admission rate, death rate |
| 19 | Taeniasis/cysticercosis | Case management (praziquantel, niclosamide, albendazole), neurological treatment, surgery | Mass drug administration (MDA) with niclosamide/praziquantel/albendazole, community awareness | Not applicable | Improved pig husbandry, pig vaccination | WASH practices, proper cooking | Prevalence, incidence, detection rate, cure rate, disability rate (neurological symptoms), surgery rate, death rate |
|  |  | Innovative and intensified disease management | Preventive chemotherapy | Vector control | Veterinary public health measures | Provision of safe WASH practices |  |
| 20 | Trachoma | Eye surgery to treat trichomatous trichiasis, TT (the S of the SAFE strategy) Azithromycin | Mass drug administration (MDA) of azithromycin and tetracycline eye ointment (WHO-recommended SAFE strategy) | Reducing breeding sites for muscid flies | Not applicable | Access to water and improved sanitation | Prevalence, incidence, detection rate, cure rate, disability rate (blindness), surgery rate |
| 21 | Yaws | Case management (azithromycin medication, intramuscular benzathine benzylpenicillin), Morges strategy, health education | Mass treatment, also known as total community treatment (TCT), involves giving oral azithromycin (single dose) | Not applicable | Not applicable | Personal hygiene | Prevalence, incidence, detection rate, cure rate, disability rate (disfiguration) |

The review will include only studies reporting primary outcomes. However, in the included studies, secondary or exploratory outcomes related to the nutrition, health, and well-being of adults and children will be analyzed to assess how NTD-related service utilization and treatment outcomes influence these outcomes in LMICs. For each of the 21 NTDs prioritized by WHO, the following framework will be examined to quantify the effect sizes: *incentive programs → increased service utilization and improved treatment outcomes → improved nutritional status and health and well-being*. Exploratory outcomes will include, but not be limited to, nutritional status, various types of malnutrition, micro- and macronutrient deficiency, anemia, cognitive development, school attendance and performance, mortality, and morbidity.

#### Study designs

The review will include randomized controlled trials (RCTs), as well as non-randomized studies with comparator groups, such as cohort, case-control, longitudinal, cross-sectional, ecological, and quasi-experimental studies. Additionally, impact evaluation results, program reports, qualitative studies, and studies employing methodologies like difference-in-difference, pre-post design, interrupted time series, regression discontinuity, instrumental variable estimation, and propensity score matching will be considered. Studies reported in English language will be considered for inclusion.

### Search strategy

A scoping search will be conducted to identify key Medical Subject Headings (MeSH) terms and other relevant search terms, which will then be combined to develop a comprehensive search strategy. Boolean operators (AND, OR, and NOT), use truncation/stemming (*), and phrase searching (“”) will be employed. The search will focus on NTDs, using genus names along with key terms, such as cash transfer, unconditional cash transfer, conditional cash transfer, micro-credit, food voucher, and incentive. A detailed preliminary search strategy is provided in Table 2.

**Table 2.** Preliminary search strategy.

| Domain | Description | Search terms/Keywords |
| --- | --- | --- |
| Population | Participants | Adults and children living in LMICs<br>Adults, young adults, young women, young men, child, infant, adolescents, teen |
| Interventions | Programs | Money, monetary, cash, financial transfer, cash transfer, payment, conditional cash transfer (CCT), unconditional cash transfer (UCT), incentive.<br>Financial, non-financial incentive programs, e.g., social protection, social program, scholarship programs, family allowance, in-kind transfer, food/medicine voucher, and micro-finance/micro-credit. |
| Outcomes | Neglected Tropical Diseases and disease groups; Core strategic interventions; NTD-related service utilization; and treatment outcomes | Neglected disease, neglected tropical disease, tropical medicine, tropical disease, Buruli, buruli ulcer, Bairnsdale ulcer, Daintree ulcer, Mossman ulcer, mycobacterium, *Mycobacterium ulcerans*, Chagas, Chagas disease, trypanosome, *Trypanosoma cruzi*, american trypanosomiasis, dengue, dengue virus, break-bone disease, break-bone fever, chikungunya, dracuncul, dracunculus nematode, dracunculiasis, guinea worm, guinea-worm, antihelminth, antiparasit, deworm, anti-parasitic, *Echinococcus*, echinococc, foodborne, trematodiasis, trematodias, clonorchiasis, opisthorchiasis, fascioliasis, paragonimiasis, human african trypanosomiasis, sleeping sickness, Leishmaniasis, kala azar, leprosy, Hansen, trypanosome, lymphatic filariasis, filaria, elephantiasis, *Wuchereria bancrofti*, brugia, onchocerciasis, rabies, river blindness, schistosoma, bilharzia, helminth, roundworm, nematode, soil transmitted helminthiasis, ascaris, whipworm, trichuris, hookworm, ancylostom, taenia, cysticercos, chlamydia, tapeworm, hookworm, *Necator americanus*, *Ancylostoma duodenale*, taenia, cysticercosis, cestod, trachoma, treponem, noma, snake bite envenoming, yaws, frambesia, bejel, pinta, parangi, mycetoma, chloroblastomysis, chromomyc, mycosis, mycos, scabies, ectoparasite, mosquito habitat, Temephos, water fleas, small liver flukes, faecal waste, faeces, lymphoedema, adenolymphangitis, hydrocele, secondary infection, agriculture, wildlife, environment, sporotrichos, feral cats, black fly habitat, larval breeding, wild mammals, veterinarians, animal handlers, rabies virus, pig husbandry, breeding sites, etc.<br>Early detection, antibiotic, Rifampicin, Clarithromycin, hygiene, wound care, BCG vaccine, Benznidazole, Nifurtimox, MDA, mass drug administration, surgery, blood screening, blood transfusion, organ transplantation, insecticide, bed nets, spraying, fluid management, wire |
|  |  | mesh, repellents, safe water practices, waste management, surveillance, cash reward scheme, voluntary reporting, Cyclops, dogs, water filtration, Albendazole, early detection by USG, deworming, deworming of dogs, sheep vaccination, handwash practices, anthelmintic medicines, Praziquantel, PAIR methods, partial hepatectomy, rehabilitation, Triclabendazole, Praziquantel, one health, WASH practices, screening, Pentamidine, Eflornithine, Fexinidazole, baits, cryotherapy, thermotherapy, liposomal Amphotericin B, pentavalent antimonials, Miltefosine, rodent control, dog vaccination, physical therapy, post-exposure prophylaxis, PEP, anti-filarial medicines, vector control, sanitation, limb care, excision, debridement, amputation, long clothes, closed-toed shoes, pet control, mouthwash, nutrition, nutrition supplement, Ivermectin, Doxycycline, eye care, pre-exposure vaccination, scabicides, snail control, antenatal care, ANC, excreta management, antivenom, ventilation, footwear, Niclosamide, neurological treatment, pig vaccination, eye surgery, Azithromycin, Tetracycline, eye ointment, SAFE strategy, muscid flies, personal hygiene.<br>Attendance at screening clinics, treatment adherence, health-seeking behavior, NTD campaign coverage, etc.<br>Prevalence, incidence, detection rate, cure rate, disability, disability rate, morbidity, morbidity rate, death rate, lymphoedema management, wound management, total cystopericystectomy, partial cystopericystectomy, surgery rate, liver damage, mental health, disfiguration, ocular morbidity, co-morbidities, kidney disease rheumatic heart disease, systemic infection, ICU, Intensive Care Unit, admission rate, blindness, |
| Study | Design | RCTs, non-randomized studies, cohort studies, longitudinal studies, case-control studies, cross-sectional studies, ecological studies, quasi-experimental studies, impact evaluation results, and program reports.<br>Difference-in-difference, pre-post design, time series, regression discontinuity, instrumental variable estimation, propensity score matching, and qualitative study. |

### Article screening process

The following databases will be searched: Medline, PubMed, Scopus, EMBASE, Web of Science, POPLINE, Google Scholar, Cochrane Collection, PsycINFO, Global Health, EconLit, Social Sciences Citation Index, International Bibliography of the Social Sciences, Knowledge Commons of Population Council, and 3ie database. Grey literature databases, including WHO websites, AlignMNH, World Bank, and UNICEF websites, will also be searched for relevant reports. Websites and online resources of universities and research centers in LMICs will be reviewed to identify working papers, dissertations and theses. Reference lists or bibliographies of included studies will also be hand-searched to include any additional relevant studies. For those studies not available in the public domain, a librarian will be contacted. Studies in English, published or available from 2000 to 2024, will be included. A search log will be maintained throughout the search process to document the database platforms or search interface used, the date the search was conducted, the search strings used, and the number of records/counts identified. The log will be updated as necessary to ensure transparency, robustness, and ease of updating during the study period.

Preliminary search documents will be stored in reference management software (EndNote/Mendeley). All the screened studies will be transferred into Microsoft Excel or Rayyan tool, a systematic review management platform (https://new.rayyan.ai/).

### Title, abstract and full-text relevancy

Two reviewers will independently screen studies based on titles and abstracts. During the initial screening phase, decisions made by each reviewer will be blinded. After the initial screening is done, the studies screened by the two reviewers will be compared and checked for discrepancies. Any discrepancies in the categorization will be resolved through consultation between the reviewers or with another reviewer. A second-level judicial review will then be conducted by a separate reviewer to perform quality checks on studies with discrepancies and those deemed ineligible. Studies deemed relevant based on titles and abstracts will undergo full-text screening for relevancy, following the same procedures used during the title and abstract screening phase. Reasons for the exclusion of papers will be recorded. The studies will be equally divided among the reviewers for relevancy screening. Data will be extracted individually by each reviewer. Any questions arising during the process will be addressed through consultation among the reviewers to reach a consensus. Eligible studies from the full-text review will be entered directly into a master data extraction spreadsheet. Figure 1 illustrates the screening process based on the PRISMA flow diagram (50).

**Figure 1.**
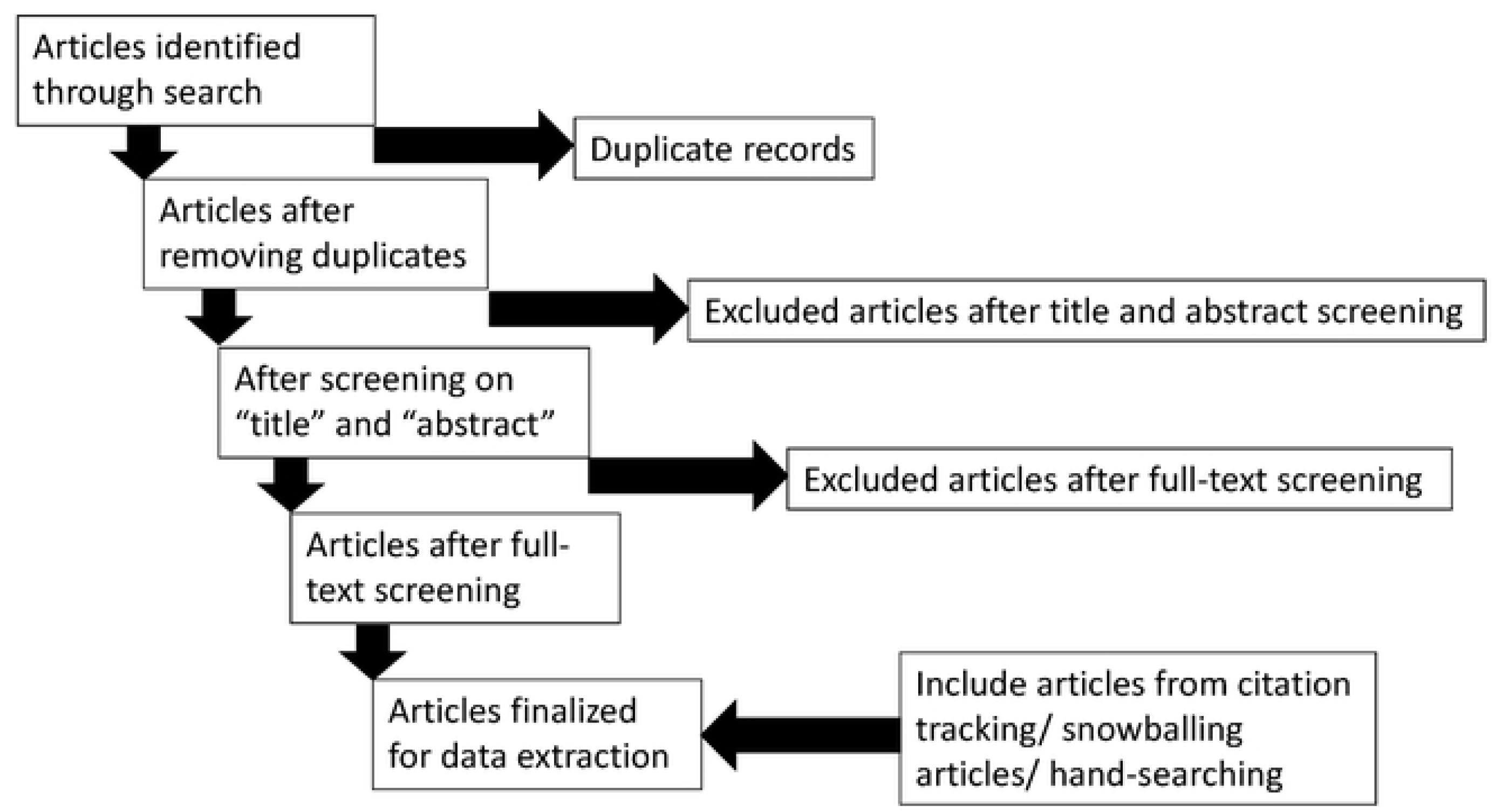
Step-by-step article screening process

### Data extraction

After the article screening process is completed, all eligible studies will be extracted into a master data extraction spreadsheet. The data extraction sheet will include the following categories: title, author(s), date of publication, type of publication (e.g., summary, synthesis, single study), number and type of included studies (if a summary or synthesis), settings and population studied, interventions implemented, outcomes measured and results, and if relevant, whether results differed among subgroups, such as gender, socioeconomic status, or ethnicity. For studies with multiple outcomes or variables, all eligible outcomes will be extracted. The data extraction form will be pilot-tested and refined based on the information extracted from a few initial studies meeting the inclusion criteria. This process will help minimize bias and improve the validity and reliability of the systematic review. Authors will be contacted through email if key information is missing or if further information or clarification is required.

### Risk of bias assessment for individual studies

The quality assessment will be done by using the Revised Cochrane Risk of Bias for randomized trials (RoB 2) (51), the Risk of Bias in Non-randomized Studies of Interventions (ROBINS-I) (52), and an adapted version of the Critical Appraisal Skills Programme (CASP) (53) for qualitative studies. The quality of each included study will be independently assessed by one reviewer, with the assessment results checked by a second reviewer. Any discrepancies or disagreements will be resolved through consultation with a third reviewer to reach a consensus.

### Anticipated number of studies to be included in the review

Based on the initial search to develop the protocol, about 20-30 studies are anticipated to be included in the review (34, 54, 55). This number might change during the actual work of the systematic review based on the search using electronic databases and grey literature.

### Data synthesis

A narrative synthesis will be produced by describing the studies. A geographical information system (GIS) map will be generated to show the frequency distribution of impact evaluations of cash transfer and incentive programs on NTD-related service utilization, treatment outcomes, nutritional status, and health and wellbeing. Interventions of cash transfers and incentive programs will be classified into different categories for different types of NTDs and their effects will be synthesized for (1) NTD-related service utilization; (2) treatment outcomes; and (3) nutritional status, and health and well-being outcomes. All the studies will be tabulated by quantitative and qualitative type and key findings will be described. Primary and secondary outcomes will be listed along with differences in effectiveness between different cash transfer and incentive programs for NTD control. The possibility of performing a meta-analysis will be explored depending on the availability of sufficient relevant studies. The ability to undertake a meta-analysis will depend on the heterogeneity of the measures, such as similarities or differences in population, intervention, outcomes, and methodologies used. If a meta-analysis is appropriate, the statistical heterogeneity of effects will be assessed using chi-square and I-square statistics. Efforts will be made to analyze the factors explaining heterogeneity through moderation analysis, including sub-group meta-analysis. For qualitative studies, the themes will be identified using a deductive approach and the findings will be used to interpret/complement the quantitative findings as part of the narrative synthesis.

The synthesis will present the factors such as program design, implementation, context, and challenges that influence the impact of cash transfer or incentive programs on NTD-related service utilization, treatment outcomes, nutrition status, and health and wellbeing. Absolute evidence gaps and synthesis gaps will also be identified that can inform the development of new studies or programs.

### Analysis of subgroups or subsets

The possibility of performing sub-group analyses will be explored. It will, however, depend on the evidence strength to summarize effects in special populations such as pregnant women, and geriatric populations, or to distinguish effect differences across different levels of groups, such as female/male, child/adolescent/adult, different geographical regions, malnourished/well-nourished, and type of disease agents (viruses, bacteria, protozoa, fungi, helminths, ectoparasites, snakebites). The effects will also be examined to determine how they differ for different covariates, program design, implementation plan, and incentive program design (type, amount, and frequency).

### Ethics and dissemination

The systematic review protocol has been reviewed and approved by two independent institutional review boards (IRBs): icddr,b (PR-24112) in Bangladesh and Population Council Institute (IORG0011824) in India. The findings of this systematic review will be presented at national and international conferences and reported in a peer-reviewed journal in accordance with the PRISMA 2020 guideline for reporting systematic reviews (50). Any amendments to this protocol during the review process will be documented in PROSPERO and reported in the final manuscript.

### Status and timeline of the study

Current status: Systematic review protocol approved by institutional review boards

Anticipated start date of the systematic review work: 1 July 2025

Anticipated title and abstract screening completion: 30 September 2025

Anticipated full-text screening completion: 31 December 2025

Anticipated data extraction completion: 31 January 2026

Anticipated first draft of the systematic review result paper: 31 March 2026

## DISCUSSION

This systematic review will be performed to critically examine and provide a comprehensive assessment of the effects of cash transfer and other financial incentive programs on NTD-related service utilization and treatment outcomes in LMICs. The economic impact of different categories of financial aid to mitigate NTDs will be assessed, which could inform policymakers on the cost-effectiveness of implementing timely intervention and other prevention or treatment efforts. Additionally, the potential link between NTD-related service utilization and health and nutrition outcomes in LMICs will be evaluated. A methodological assessment of the published literature will also be performed and the findings from the published works will be compared with those of other similar reviews. The findings from this systematic review will be compared and/or contrasted with similar published papers and the strength of evidence will be examined. Conclusions will be drawn from this systematic review highlighting the effects of several financial incentive programs on service utilization and treatment outcomes related to NTDs as well as their overall impact on health and nutrition in LMICs. Limitations of the review will be discussed in detail. The public health implications of all the findings of this systematic review will also be comprehensively discussed.

## CONCLUSION

The proposed systematic review will synthesize evidence generated by studies over a span of 25 years, critically assess the strengths and limitations of existing studies on NTDs, and evaluate the possible link between cash transfer and financial incentive programs with NTD. Rigorous methodology and robust article screening process have the potential to ensure a comprehensive, up-to-date, and unbiased evidence synthesis of the effectiveness of cash transfer and incentive programs on service utilization and treatment outcomes related to NTDs and their impact on health and nutrition in LMICs

## Data Availability

No datasets were generated or analysed during the current study. All relevant data from this study will be made available upon study completion.

## Funding

This study has been funded by the Children’s Investment Fund Foundation (CIFF).

## Disclaimer

CIFF was not involved in the design of the study, data collection and analysis, decisions regarding publication, or the preparation of the manuscript.

## Competing interests

The authors declare that they have no competing interests.

## Abbreviations

CASP: Critical Appraisal Skills Programme
CCT: Conditional Cash Transfer
LMICs: Low-and-Middle Income Countries
NTD: Neglected Tropical Disease
WHO: World Health Organization

## CONTRIBUTORS/ACKNOWLEDGEMENTS

SMTH, RD, AAN, and AH initiated the protocol and conceptualized the research plan for the proposed systematic review. SMTH, RD, AAN, and SM finalized the methodology. SMTH, RD, AAN, SG, and PC wrote the manuscript. AH, SM, STR, NK, DM, and TA critically reviewed it for important intellectual content. All authors read and approved of the final manuscript.

## SUPPORTING INFORMATION

A PRISMA-P checklist for this protocol is provided in Supplemental File 1.

## Notes

### Competing Interest Statement

The authors have declared no competing interest.

### Funding Statement

Yes

### Author Declarations

The systematic review protocol has been reviewed and approved by two independent institutional review boards (IRBs): icddr,b (PR-24112) in Bangladesh and Population Council Institute (IORG0011824) in India.

